# The health effects in the US of quarantine policies based on predicted individual risk of severe COVID-19 outcomes

**DOI:** 10.1101/2021.03.21.21254065

**Authors:** Sam Lovick, Gillian S. Dite, Richard Allman

**Author notes:** **Corresponding Author:** Richard Allman, Genetic Technologies Ltd, 60–66 Hanover St, Fitzroy VIC 3065, Australia.

## Abstract

**Background:** Social distancing, testing and public health measures are the principal protections against COVID-19 in the US. Social distancing based on an accurate assessment of the individual risk of severe outcomes could reduce harm even as infection rates accelerate.

**Methods:** An SEIR dynamic transmission model of COVID-19 was created to simulate the disease in the US after October 2020. The model comprised 8 age groups with US-specific contact rates and low- and high-risk sub-groups defined in terms of the risk of a severe outcome determined by relevant comorbidities and a genetic test. Monte Carlo analysis was used to compare quarantine measures applied to at risk persons identified with and without the genetic test.

**Results:** Under the piecemeal social distancing measures currently in place, absent a vaccine the US can expect 114 million symptomatic infections, 4.8 million hospitalisations and 262,000 COVID-19 related deaths. Social distancing based solely on comorbidities with 80% compliance reduces symptomatic infections by between 1.2 and 2.2 million, hospitalisations by between 1.2 and 1.3 million, and deaths by between 71,800 and 80,900. Refining the definition of at risk using a test of single-nucleotide polymorphisms further reduces symptomatic infections by 1.0 to 1.2 million, hospitalisations by 0.4 million and deaths by between 20,500 and 24,100.

**Conclusions:** Models are now available that can accurately predict the likelihood of severe COVID-19 outcomes based on age, sex, comorbidities and polygenetic testing. Quarantine based on risk of severe outcomes could substantially reduce pandemic harm, even when infection rates outside of quarantine are high.

## Introduction

The current COVID-19 pandemic is causing enormous harm globally from both health and economic perspectives. In some population groups, up to 30% of those infected will experience severe symptoms requiring hospitalisation; some of those will require intensive care and some will die.^1,2^

Globally, public health responses have been aimed at reducing spread and protecting high risk groups such as the elderly in residential care. Pending a vaccine, governments have sought to reduce community transmission through measures such as rapid and high volume testing, track and trace, quarantine, social distancing, mask wearing, curtailing non-essential services and travel restrictions.

Extensive social distancing including travel constraints has been shown to control COVID-19 spread in practice.^3^ Disease modelling has also identified the importance of these measures on disease spread and pandemic harm.^4^ However, the economic and social impacts of these interventions have been substantial, with considerable damage to local economies.^5^ There have also been allied health effects such as increases in reported mental health diagnoses^6^ and decreases in cancer screening heralding likely increases in future cancer deaths.^7^

Most of these social distancing regimes have focused on measures applied broadly across the population. While some states within the US have temporarily imposed broad-based measure, the US as a whole appears reluctant to impose the stringent measures that have been a feature of Australia’s and some European countries’ responses to COVID-19.

Infection rates in northern hemisphere countries in Europe and North America have increased markedly as initial measures that were successful in curbing infection rates were relaxed.^8^ In the US, close to 10% of hospital beds were occupied with COVID-19 patients before the July 2020 second peak in infections. In the worst affected states, more than 25% of beds were occupied with COVID-19 with similarly high ICU bed occupancy.^9^ US infection rates in November 2020 are considerably higher, resulting in concomitant stresses on hospital care. One of the policy imperatives is therefore to ensure that rising infection rates do not overwhelm hospital capacity.

There is growing awareness of the factors that affect patient outcomes after infection. Age, gender and comorbidities have been identified as factors affecting the risk of a severe outcome (one requiring hospitalisation).^10^ Several epidemiological studies have sought to quantify the importance of these factors.^11,12^ To more accurately identify those at risk of a severe outcome, Dite et al examined a UK database of health and genomic data combing age, sex, comorbidities and a screening test based on 62 single-nucleotide polymorphisms (‘the SNP test’).^13^ The SNP test markedly improved identification of those at risk.

We used disease modelling to examine the health effects of social distancing in the US based on quarantining those individuals most likely to have a severe COVID-19 outcome upon infection. The potential benefit of such an approach is that for a given level of COVID-19 infections, it results in fewer hospitalisations, ICU admissions and deaths.

## Methods

### The disease model

A conventional four pool (susceptible, exposed, infectious, recovered — SEIR) dynamic transmission model developed to simulate influenza pandemics written in R using the EpiModel^14^ library was adapted to simulate COVID-19. Disease spread within the population is based on rates of contact between 8 defined age-groups across four activity areas: home, work, school and leisure/other derived after Mossong et al^15^ (undertaken as part of the European Union POLYMOD initiative^16^), extended to 136 countries including the US based on the demographic and economic characteristics of each country.^17^ Daily contact rates in the US by age for school, work, home and all other locations are shown in Supplementary Table 1.

Disease characteristics are defined in terms of naïve population R_0_ values, symptomatic and asymptomatic proportions, differential rates of spread from symptomatic and asymptomatic infections, case fatality rate for symptomatic infections, transitional probabilities for different outcomes (primary care consultation, hospitalisation, critical care and death), and typical duration of illness, hospitalisation and critical care to determine overall demand for health care services. These were largely based on Ferguson et al.^4^

The model includes details of health care capabilities in each country. These include hospital beds and critical care beds.^9^ The costs of each of these resources are also included.^18,19^ The model can therefore estimate utilisation of these resources under different intervention strategies. The model also estimates health burden in terms of the value of saved quality of life years (QALYs) assuming QALYs lost from symptomatic COVID-19 infections reflect QALYs lost from symptomatic influenza-like infections.^20^ The model assumes a $50,000 cost for each lost QALY.

The model uses second-order Monte Carlo analysis to represent uncertain factors such as R_0_, case fatality rate (CFR), and duration of infection with Latin hypercube sampling to ensure representative numbers of cases. The core modelling assumptions are set out in Table 1 and Table 2.

**Table 1.**
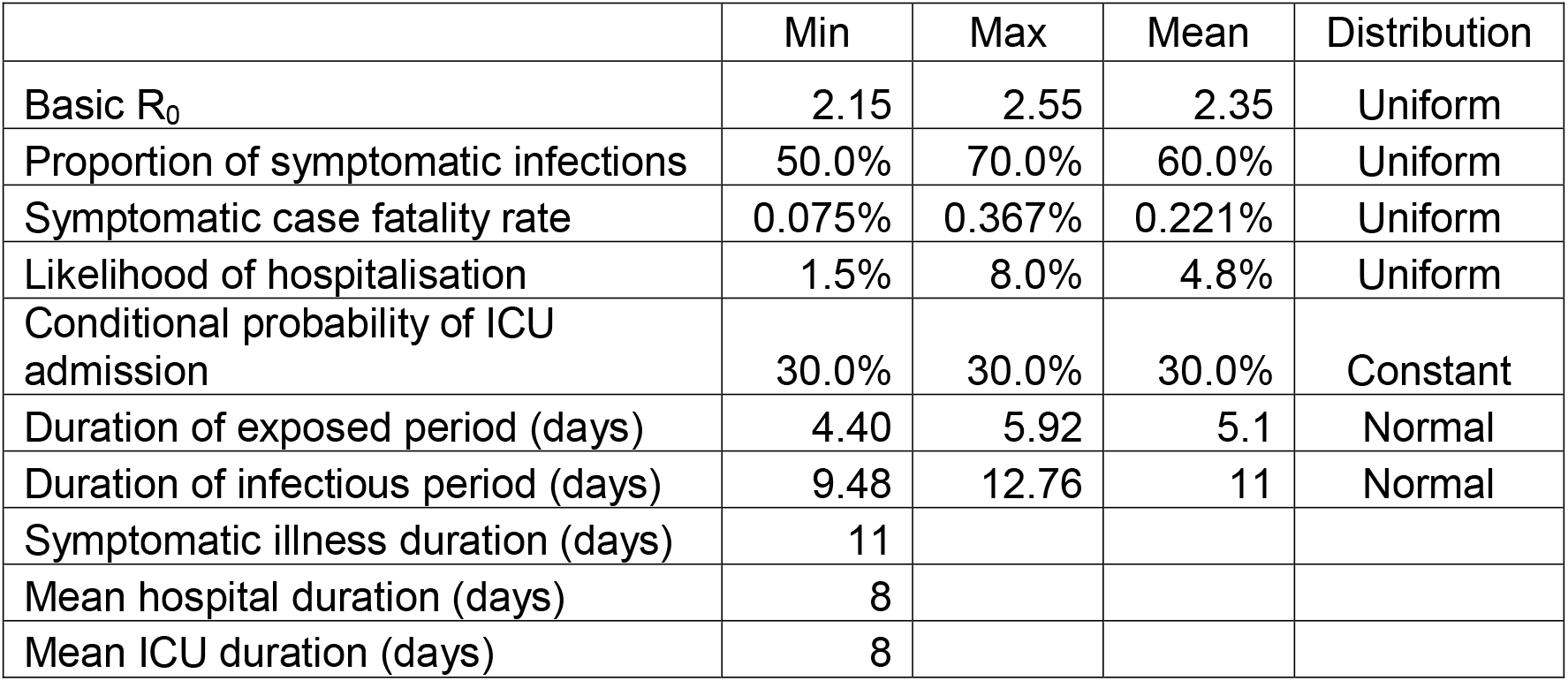
Core disease modelling assumptions absent interventions

|  | Min | Max | Mean | Distribution |
| --- | --- | --- | --- | --- |
| Basic $R_0$ | 2.15 | 2.55 | 2.35 | Uniform |
| Proportion of symptomatic infections | 50.0% | 70.0% | 60.0% | Uniform |
| Symptomatic case fatality rate | 0.075% | 0.367% | 0.221% | Uniform |
| Likelihood of hospitalisation | 1.5% | 8.0% | 4.8% | Uniform |
| Conditional probability of ICU admission | 30.0% | 30.0% | 30.0% | Constant |
| Duration of exposed period (days) | 4.40 | 5.92 | 5.1 | Normal |
| Duration of infectious period (days) | 9.48 | 12.76 | 11 | Normal |
| Symptomatic illness duration (days) | 11 |  |  |  |
| Mean hospital duration (days) | 8 |  |  |  |
| Mean ICU duration (days) | 8 |  |  |  |

**Table 2.** Relative risk of hospitalisation and death by age

| Age (years) | Relative risk of hospitalisation/death |
| --- | --- |
| 0–4 | 1% |
| 5–9 | 1% |
| 10–14 | 4% |
| 15–19 | 4% |
| 20–24 | 16% |
| 25–29 | 16% |
| 30–34 | 41% |
| 35–39 | 41% |
| 40–44 | 64% |
| 45–49 | 64% |
| 50–54 | 132% |
| 55–59 | 132% |
| 60–64 | 215% |
| 65–69 | 215% |
| 70–74 | 315% |
| 75–79 | 315% |
| 80+ | 354% |

### Social distancing assumptions

The model was adapted to include a number of different social distancing policies after Ferguson et al in the UK in March 2020.^4^ They are: school closures; work from home requirements; age-based quarantine; case quarantine; household quarantine where there is an infection in the household; and broad-based social distancing comprising restrictions on activities outside the home, school and workplace. In addition, the model was adapted to allow quarantine (with a specified rates of compliance) of adult individuals determined to be at high risk of severe outcomes under the Dite model.^13^ The Dite model is summarised in Supplementary Table 2.

These policies can be used singly or in combination. Appropriate adjustments are made to the rates of contact and risk of infection for each of the social distancing measures. Start-week and end-week for each measure can be specified. In addition, measures can be endogenously applied by the model based on the instant underlying COVID-19 mortality rate in the population as a whole.

In this modelling, school closure programs and work from home social distancing were applied when the daily COVID-19 mortality rate rose above 0.5 deaths per 100,000 population per day. We also assumed that 50% of symptomatic infections are identified from testing sufficiently early for effective quarantine of those individuals to prevent further spread. This quarantine measure was adopted once the daily COVID-19 mortality rate rose above 0.1 deaths per 100,000 population per day. Cases were quarantined for 14 days. This established the base for comparison of different social distancing measures based on at risk status.

Quarantine based on at risk status, when in place, was also imposed when the daily COVID-19 mortality rate rose above 0.1 deaths per 100,000 population per day. Social distancing parameters are summarised in Supplementary Table 3.

### Characterisation of at risk individuals

The proportion of the population by age and sex with comorbidities was gathered from a number of sources. Where used: body mass index^21^; smoking^22^; heart disease^23–25^; hyperlipidaemia^26^; hypertension^27^; diabetes^28^; COPD^29^ and asthma^30^; kidney disease^31^; cerebrovascular^25^; kidney disease^32^; and cancer^33^. The comorbidities were assumed to be independent, an assumption that could be relaxed in future simulations. Relevant comorbidity rates by age and sex in the base population are summarised in Supplementary Table 4.

The odds ratios for each of the factors were taken from the model developed by Dite et al^13^ with only those factors with an odds ratio p value of 0.001 or below included in the analysis. The proportion of at risk in each age group for the purposes of the disease modelling was set as the proportion of the cohort exhibiting an odds ratio above of 2.5 and 3.5 of a severe outcome post infection. These proportions were determined with and without the SNP test. Rates of at risk within the population are summarised in Supplementary Tables 5 and 6 for the 2.5 and 3.5 odds ratio threshold respectively.

Population sizes by age was based on US census data^34^. Age cohorts were specified for years 0 to 10, 10 to 18, 18 to 30, deciles to 70, and 70 years and above. Social distancing measures for high-risk individuals applied only to adults 18 years and above.

## Results

Left to run its course from October 2020 with no social distancing measures in place, the model predicts 232.6 m new infections (SD 3.2 m), 139.9 m symptomatic infections (SD 4.7 m), 5.7 m hospitalisations (SD 0.7 m) and 370,100 deaths (SD 48,500).

The profile of disease spread absent social distancing appears more rapid than is currently observed (notwithstanding uncertainty over actual rather than observed infection rates) and the US is putting some social distancing measures in place albeit with wide variation in timing and extent across the states. With limited social distancing comprising school closure, home-based work and infected case quarantine endogenously triggered based on mortality rate, the model predicts 190.5 m infections (SD 4.1 m), 114.3 m symptomatic infections (SD 3.7 m), 4.8 m hospitalisations (SD 0.6 m) and 262,100 deaths (SD 34,100). The predicted weekly profile of COVID-19 spread with and without limited social distancing is shown in Figure 1a. Weekly symptomatic COVID-19 infections are shown in Figure 1b and weekly predicted deaths in Figure 1c.

**Figure 1a.**
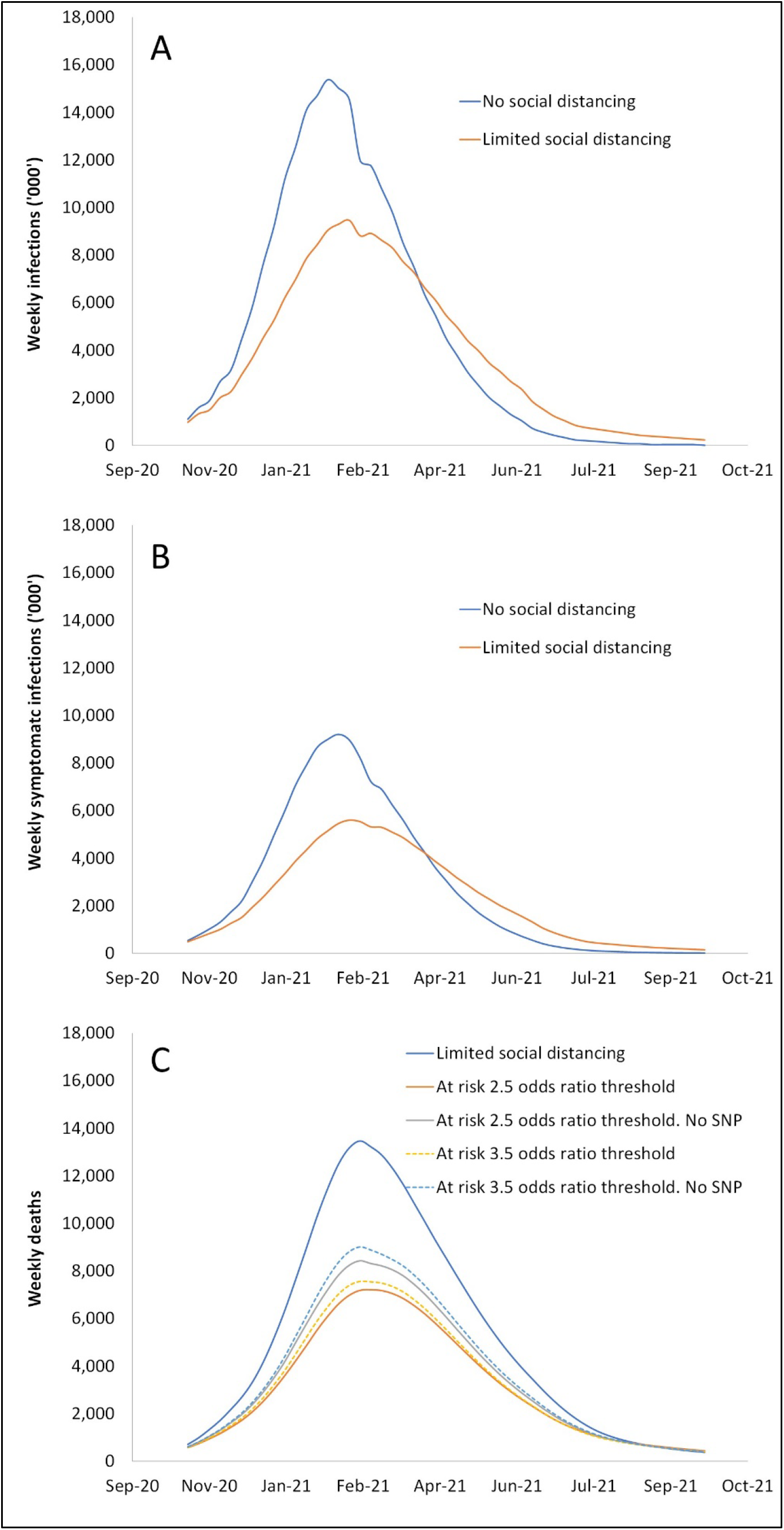
Predicted weekly cases of COVID-19 (symptomatic and asymptomatic) with and without limited social distancing without a vaccine. **1b**. Predicted weekly symptomatic COVID-19 cases (symptomatic and asymptomatic) with and without limited social distancing without a vaccine. **1c**. Predicted weekly deaths with limited social distancing and quarantine of at risk individuals using different odds ratio thresholds, with and without SNP test.

Table 3 summarises health outcomes with and without at risk social distancing assuming either an 80% compliance or 50% compliance with the social distancing measures. Where 80% compliance is achieved, at risk social distancing makes a small 1% to 3% difference to the number of infections and symptomatic cases when compared to the limited social distancing case. At risk social distancing using a 2.5 odds ratio threshold but without the SNP test results in 172,000 deaths (SD 23,500) compared to 274,000 (SD 35,000) from limited social distancing alone, a reduction of 87,200 deaths. Inclusion of the SNP test to identify at risk individuals results in a further reduction of 23,200 deaths, a 27% increase in lives saved. Inclusion of the SNP test results in a similar reduction in deaths using the higher 3.5 odds ratio threshold for classifying at risk individuals. Reductions in hospitalisations show a similar pattern.

**Table 3.**
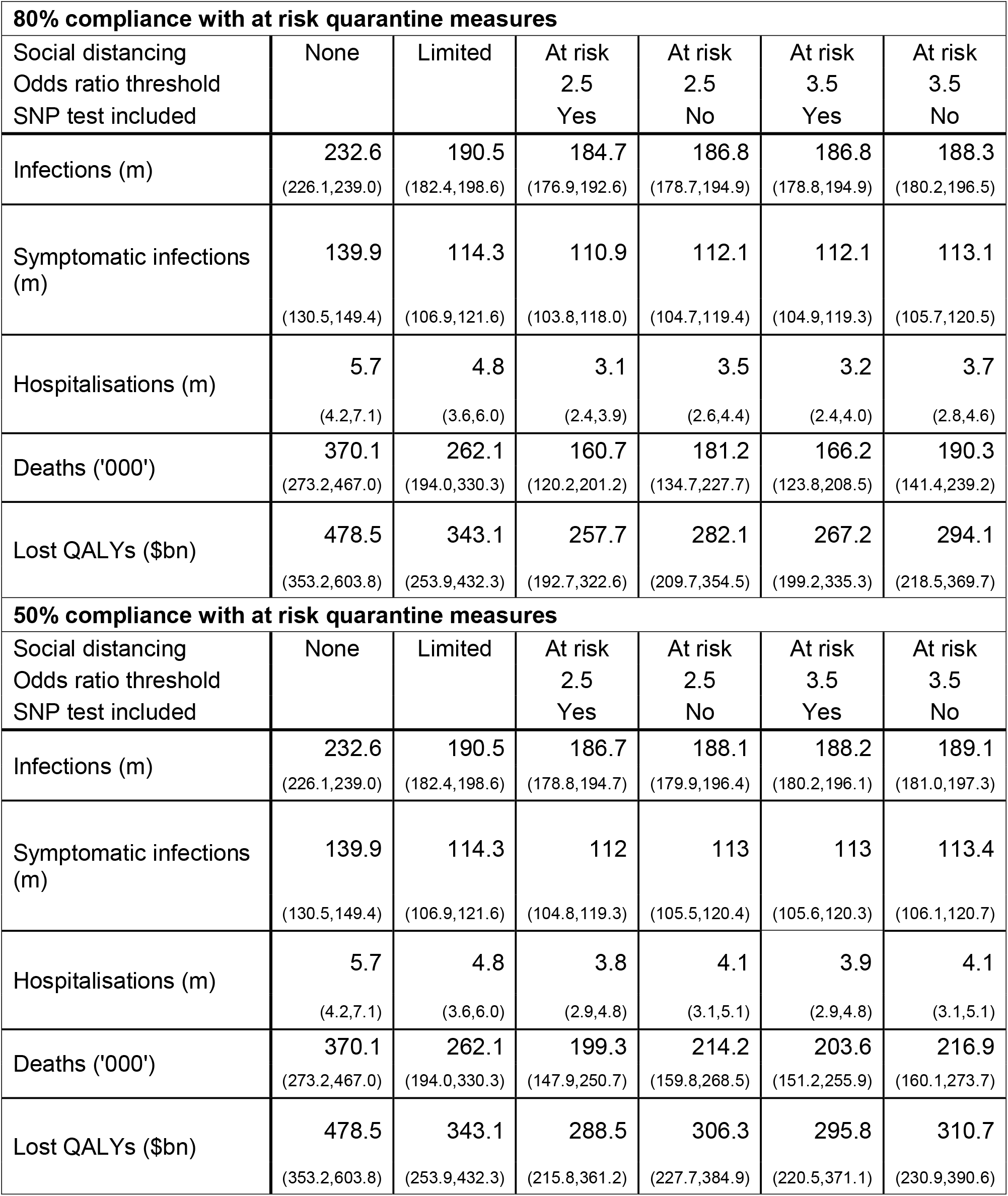

Where a lower compliance rate of 50% is achieved, at risk social distancing without the SNP test reduces deaths by 52,300. Inclusion of the SNP test results in a further reduction in deaths of 14,900, a 28% increase in lives saved. Figure 3 shows the profile of deaths during the pandemic with limited social distancing, and under all at risk-based social distancing scenarios, with and without the SNP test.

Table 3 shows that at risk-based social distancing and limited social distancing deliver similar reductions in infections and symptomatic infections. At risk-based social distancing is much more effective at reducing severe outcomes. The largest effect on severe outcomes is in the older cohorts which exhibit the highest odds ratio of severe outcomes, as shown in Supplementary Tables 7 and 8, Supplementary Figures 1 and 2.

## Discussion

Countries around the globe are grappling with the complex task of reducing the health burden from the COVID-19 pandemic. This typically involves a trade-off between social distancing and public health measures aimed at reducing infections to deliver a concomitant reduction in hospitalisations and deaths, against minimising the economic consequences that arise from restrictions of activity.

After some initial success, most European countries have seen dramatic rises in infections following relaxation of initial protective measures. Most have had to re-impose strict economy-wide social restrictions to curb infection rates. The US has, in most regions, imposed less stringent social distancing measures, which is reflected in high and rising infection rates in the fall of 2020 approaching winter.

There is growing evidence^10–13^ showing an identifiable group with a high risk of severe outcomes (hospitalisation, ICU admission and death). This analysis suggests that quarantine or social distancing applied to accurately identified “at risk” individuals could reduce these severe outcomes by a much greater amount than alternative social distancing measures covering a similar but broader-based proportion of the population.

This analysis also shows that improving the accuracy of identification of those at risk, in this case using an SNP test, markedly improves the effectiveness of an at risk-based social distancing regime. These simulations indicate that 26% of the population would need to be SNP tested when the 2.5 odds ratio screen is used, 15% when the 3.5 odds ratio screen is used. Given these testing rates, each test would generate between $285 and $360 in the value of saved QALYs.

In this analysis we have not sought to examine the practicality of an at risk social distancing policy. However, we already know that where there is concentration of at risk individuals, notably in care homes, specific safeguard to protect against sources of COVID-19 are advisable.^35^ Such specific safeguards are, in effect, a form of at risk social distancing.

This analysis indicates that, where governments choose minimal levels of social distancing to safeguard the economy, social distancing targeting those at most risk will be particularly valuable in minimising the burden of disease, not least because retirees are a large proportion of that group for COVID-19. Improving the accuracy of identification through, for example, a low cost genetic test, increases this value. And while this analysis examines the US as a whole, the conclusions are equally applicable at the regional or community level.

To date, there has been only limited study of the use of targeted quarantine measures as a means of limiting pandemic harm, whether in terms of burden of disease or economic harm. In part, this reflects the difficulty in identifying and the subsequent costs of quarantining individuals. The value of this study is an assessment of genetic testing to better identify at risk individuals, which could change the relative costs and benefits of targeted quarantine.

The evidence indicates that single-nucleotide polymorphism testing, combined with age, sex and comorbidities, can accurately identify those at risk of severe COVID-19 outcomes, making quarantine of those individuals within communities, whether small or large, a more viable response to the pandemic.

## Data Availability

N/A

## Footnote

The modelling was undertaken in October 2020 using data on the social distancing measures that States had implemented up to that time as the base social distancing assumptions. Subsequently and after this paper was drafted, there was an effective relaxation of social distancing measures in the US as COVID spread to US regions that adopted less stringent measures and following Thanksgiving travel. In combination, these resulted in outcomes that were materially worse than forecast. Had these events not taken place, outcomes in the US would have been in line with this forecast.

## Ethical Considerations

Ethics approval not required as the research is a computer modelling study.

## Authorship Statement

RA and SL conceived the study concept and design. GSD and SL contributed to acquisition of data. SL conducted the data analysis. SL drafted the first version of the manuscript. All authors contributed to interpretation of data and critical revision of the manuscript.

## Supplementary Tables

**Supplementary Table 1.**
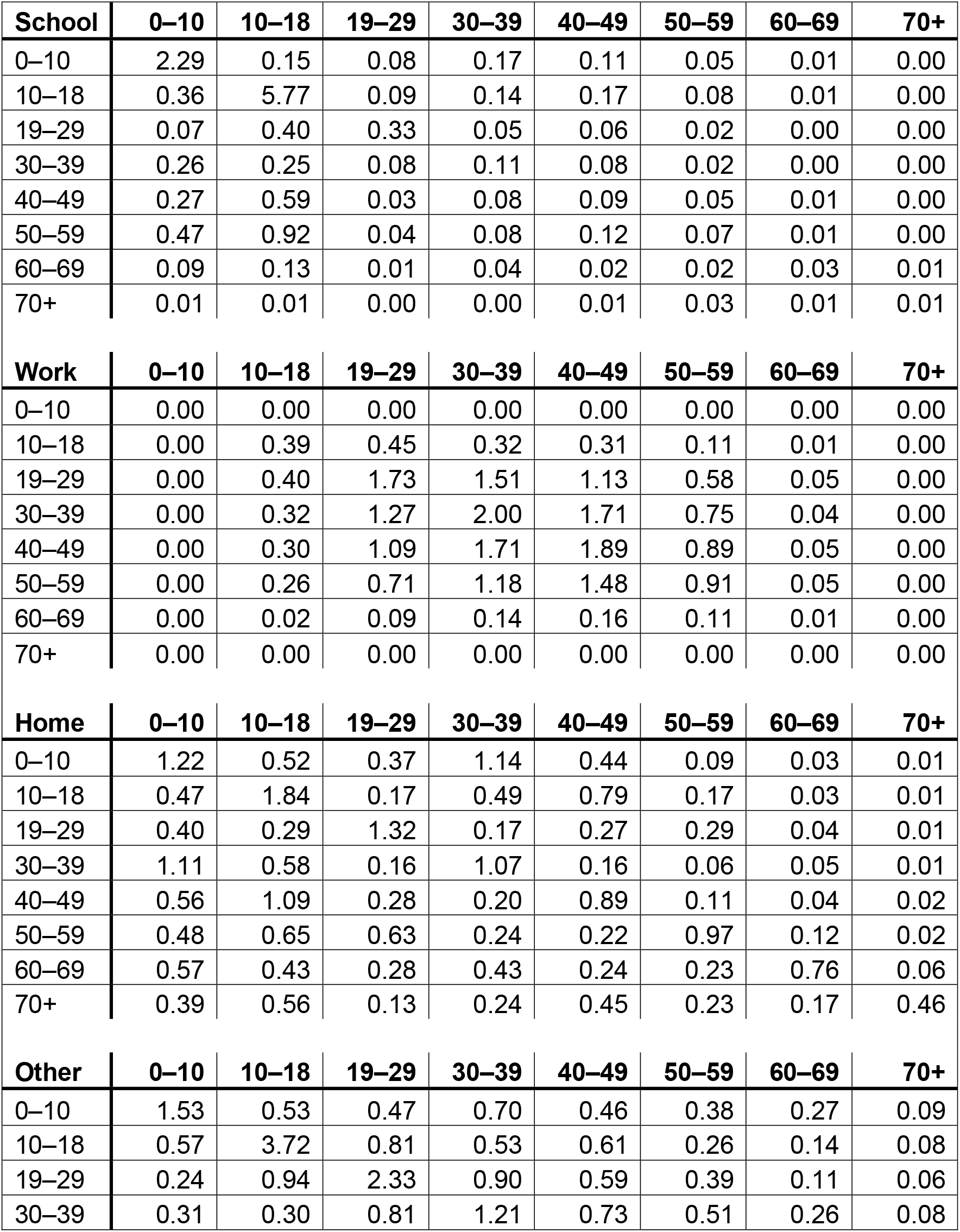
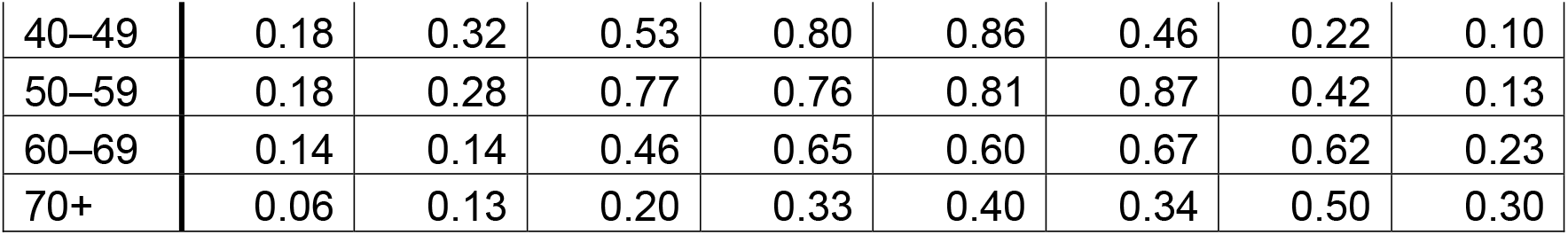
Daily contact rates

| **School** | **0–10** | **10–18** | **19–29** | **30–39** | **40–49** | **50–59** | **60–69** | **70+** |
| --- | --- | --- | --- | --- | --- | --- | --- | --- |
| 0–10 | 2.29 | 0.15 | 0.08 | 0.17 | 0.11 | 0.05 | 0.01 | 0.00 |
| 10–18 | 0.36 | 5.77 | 0.09 | 0.14 | 0.17 | 0.08 | 0.01 | 0.00 |
| 19–29 | 0.07 | 0.40 | 0.33 | 0.05 | 0.06 | 0.02 | 0.00 | 0.00 |
| 30–39 | 0.26 | 0.25 | 0.08 | 0.11 | 0.08 | 0.02 | 0.00 | 0.00 |
| 40–49 | 0.27 | 0.59 | 0.03 | 0.08 | 0.09 | 0.05 | 0.01 | 0.00 |
| 50–59 | 0.47 | 0.92 | 0.04 | 0.08 | 0.12 | 0.07 | 0.01 | 0.00 |
| 60–69 | 0.09 | 0.13 | 0.01 | 0.04 | 0.02 | 0.02 | 0.03 | 0.01 |
| 70+ | 0.01 | 0.01 | 0.00 | 0.00 | 0.01 | 0.03 | 0.01 | 0.01 |
| **Work** | **0–10** | **10–18** | **19–29** | **30–39** | **40–49** | **50–59** | **60–69** | **70+** |
| 0–10 | 0.00 | 0.00 | 0.00 | 0.00 | 0.00 | 0.00 | 0.00 | 0.00 |
| 10–18 | 0.00 | 0.39 | 0.45 | 0.32 | 0.31 | 0.11 | 0.01 | 0.00 |
| 19–29 | 0.00 | 0.40 | 1.73 | 1.51 | 1.13 | 0.58 | 0.05 | 0.00 |
| 30–39 | 0.00 | 0.32 | 1.27 | 2.00 | 1.71 | 0.75 | 0.04 | 0.00 |
| 40–49 | 0.00 | 0.30 | 1.09 | 1.71 | 1.89 | 0.89 | 0.05 | 0.00 |
| 50–59 | 0.00 | 0.26 | 0.71 | 1.18 | 1.48 | 0.91 | 0.05 | 0.00 |
| 60–69 | 0.00 | 0.02 | 0.09 | 0.14 | 0.16 | 0.11 | 0.01 | 0.00 |
| 70+ | 0.00 | 0.00 | 0.00 | 0.00 | 0.00 | 0.00 | 0.00 | 0.00 |
| **Home** | **0–10** | **10–18** | **19–29** | **30–39** | **40–49** | **50–59** | **60–69** | **70+** |
| 0–10 | 1.22 | 0.52 | 0.37 | 1.14 | 0.44 | 0.09 | 0.03 | 0.01 |
| 10–18 | 0.47 | 1.84 | 0.17 | 0.49 | 0.79 | 0.17 | 0.03 | 0.01 |
| 19–29 | 0.40 | 0.29 | 1.32 | 0.17 | 0.27 | 0.29 | 0.04 | 0.01 |
| 30–39 | 1.11 | 0.58 | 0.16 | 1.07 | 0.16 | 0.06 | 0.05 | 0.01 |
| 40–49 | 0.56 | 1.09 | 0.28 | 0.20 | 0.89 | 0.11 | 0.04 | 0.02 |
| 50–59 | 0.48 | 0.65 | 0.63 | 0.24 | 0.22 | 0.97 | 0.12 | 0.02 |
| 60–69 | 0.57 | 0.43 | 0.28 | 0.43 | 0.24 | 0.23 | 0.76 | 0.06 |
| 70+ | 0.39 | 0.56 | 0.13 | 0.24 | 0.45 | 0.23 | 0.17 | 0.46 |
| **Other** | **0–10** | **10–18** | **19–29** | **30–39** | **40–49** | **50–59** | **60–69** | **70+** |
| 0–10 | 1.53 | 0.53 | 0.47 | 0.70 | 0.46 | 0.38 | 0.27 | 0.09 |
| 10–18 | 0.57 | 3.72 | 0.81 | 0.53 | 0.61 | 0.26 | 0.14 | 0.08 |
| 19–29 | 0.24 | 0.94 | 2.33 | 0.90 | 0.59 | 0.39 | 0.11 | 0.06 |
| 30–39 | 0.31 | 0.30 | 0.81 | 1.21 | 0.73 | 0.51 | 0.26 | 0.08 |
| 40–49 | 0.18 | 0.32 | 0.53 | 0.80 | 0.86 | 0.46 | 0.22 | 0.10 |
| 50–59 | 0.18 | 0.28 | 0.77 | 0.76 | 0.81 | 0.87 | 0.42 | 0.13 |
| 60–69 | 0.14 | 0.14 | 0.46 | 0.65 | 0.60 | 0.67 | 0.62 | 0.23 |
| 70+ | 0.06 | 0.13 | 0.20 | 0.33 | 0.40 | 0.34 | 0.50 | 0.30 |

**Supplementary Table 2.**
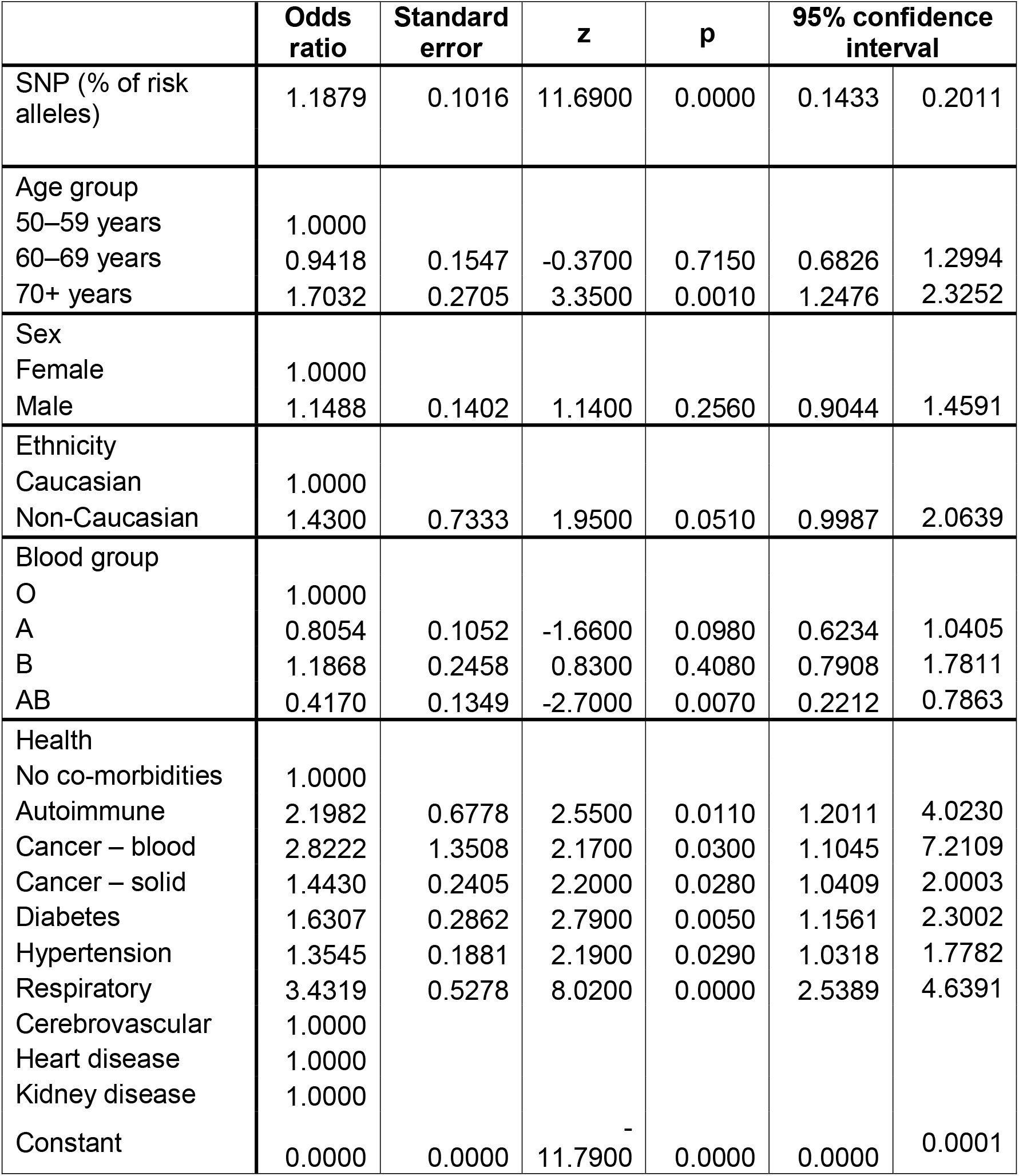
Dite model of risk of severe outcomes incorporating SNP test

**Supplementary Table 3.** Social distancing parameters

| Measure | Impact on rates of contact |  |  |  | Mortality threshold<br>(deaths/100,000/day) |
| --- | --- | --- | --- | --- | --- |
|  | School | Work | Home | Other |  |
| School closures | -80% | 0% | 20% | 0% | 0.5 |
| Work from home | 0% | -20% | 20% | 0% | 0.5 |
| Case isolation | -10% | 25% | 25% | -80% | 0.1 |
| Quarantine at risk | 5% | -100% | 0% | -100% | 0.1 (when selected) |

**Supplementary Table 4.** Proportion of adult population with relevant comorbidities

|  | Age band (years) | 18–29 | 30–39 | 40–49 | 50–59 | 60–69 | 70+ |
| --- | --- | --- | --- | --- | --- | --- | --- |
| Diabetes | Male | 0.6% | 2.6% | 5.4% | 9.1% | 13.8% | 22.7% |
|  | Female | 0.9% | 1.8% | 3.5% | 6.2% | 9.8% | 16.9% |
| Autoimmune |  | 0.5% | 0.5% | 1.5% | 4.0% | 4.0% | 6.5% |
| Respiratory |  | 2.5% | 3.0% | 3.5% | 6.0% | 8.0% | 9.5% |
| Hypertension | Male | 8.2% | 17.0% | 24.7% | 32.4% | 40.0% | 51.5% |
|  | Female | 1.2% | 10.3% | 18.2% | 26.1% | 34.0% | 45.9% |
| Cancer - solid |  | 0.3% | 0.8% | 2.4% | 4.8% | 7.9% | 16.0% |
| Cancer - blood |  | 0.1% | 0.2% | 0.3% | 0.4% | 0.5% | 0.9% |
| Cerebrovascular | Male | 0.7% | 0.7% | 3.9% | 3.9% | 6.7% | 10.9% |
|  | Female | 0.5% | 0.5% | 3.4% | 3.4% | 5.5% | 11.3% |
| Heart disease | Male | 3.6% | 3.6% | 10.2% | 16.3% | 28.8% | 42.0% |
|  | Female | 4.3% | 4.3% | 9.0% | 13.1% | 17.7% | 31.2% |
| Kidney disease | Male | 3.2% | 4.7% | 7.9% | 10.5% | 19.9% | 31.9% |
|  | Female | 4.0% | 6.0% | 10.0% | 13.0% | 25.0% | 40.0% |

**Supplementary Table 5.** Characteristics of at risk persons (odds ratio threshold 2.5)

| Age<br>(years) | Comorbidities and SNP test |  |  | Comorbidities only |  |  |
| --- | --- | --- | --- | --- | --- | --- |
|  | % at<br>risk | Average<br>number of<br>comorbidities | Odds<br>ratio<br>above<br>threshold | % at<br>risk | Average<br>number of<br>comorbidities | Odds<br>ratio<br>above<br>threshold |
| 18–29 | 0.2% | 1.23 | 3.39 | 0.0% | 1.00 | 1.00 |
| 30–39 | 1.1% | 1.28 | 3.61 | 0.1% | 2.33 | 3.22 |
| 40–49 | 5.4% | 1.37 | 4.02 | 1.3% | 2.20 | 3.42 |
| 50–59 | 16.6% | 1.53 | 4.75 | 10.1% | 1.98 | 3.88 |
| 60–69 | 19.3% | 2.02 | 5.03 | 12.3% | 2.43 | 4.24 |
| 70+ | 49.0% | 2.24 | 7.07 | 44.9% | 2.49 | 5.49 |

**Supplementary Table 6.** Characteristics of at risk persons (odds ratio threshold 3.5)

| Age<br>(years) | Comorbidities and SNP test |  |  | Comorbidities only |  |  |
| --- | --- | --- | --- | --- | --- | --- |
|  | % at<br>risk | Average<br>number of<br>comorbidities | Odds ratio<br>above<br>threshold | % at<br>risk | Average<br>number of<br>comorbidities | Odds ratio<br>above<br>threshold |
| 18–29 | 0.0% | 1.80 | 5.14 | 0.0% | 1.00 | 1.00 |
| 30–39 | 0.4% | 1.34 | 4.78 | 0.0% | 2.50 | 5.16 |
| 40–49 | 2.3% | 1.53 | 5.46 | 0.4% | 2.59 | 4.57 |
| 50–59 | 9.2% | 1.74 | 6.23 | 4.7% | 2.32 | 5.04 |
| 60–69 | 11.2% | 2.20 | 6.54 | 5.9% | 2.73 | 5.58 |
| 70+ | 35.1% | 2.41 | 8.69 | 29.5% | 2.69 | 6.88 |

**Supplementary Table 7.** Deaths by age, 80% compliance with at risk quarantine measures

|  |  |  |  |  |  |  |
| --- | --- | --- | --- | --- | --- | --- |
| Social distancing | None | Limited | At risk | At risk | At risk | At risk |
| Odds ratio threshold |  |  | 2.5 | 2.5 | 3.5 | 3.5 |
| SNP test included |  |  | Yes | No | Yes | No |
| 18–30 years | 10,888 | 7,629 | 7,177 | 7,261 | 7,214 | 7,338 |
| 30–40 years | 24,580 | 17,175 | 15,795 | 16,295 | 16,016 | 16,510 |
| 40–50 years | 36,189 | 25,149 | 20,888 | 23,264 | 21,918 | 23,883 |
| 50–60 years | 78,903 | 54,331 | 34,923 | 41,608 | 37,897 | 45,348 |
| 60–70 years | 83,625 | 57,901 | 35,143 | 41,848 | 37,907 | 45,870 |
| 70–85 years | 133,235 | 98,062 | 45,007 | 49,164 | 43,431 | 49,552 |

**Supplementary Table 8.** Deaths by age, 50% compliance with at risk quarantine measures

|  |  |  |  |  |  |  |
| --- | --- | --- | --- | --- | --- | --- |
| Social distancing | None | Limited | At risk | At risk | At risk | At risk |
| Odds ratio threshold |  |  | 2.5 | 2.5 | 3.5 | 3.5 |
| SNP test included |  |  | Yes | No | Yes | No |
| 18–30 years | 10,888 | 7,629 | 7,279 | 7,411 | 7,351 | 7,384 |
| 30–40 years | 24,580 | 17,175 | 16,158 | 16,653 | 16,407 | 16,617 |
| 40–50 years | 36,189 | 25,149 | 22,254 | 24,003 | 23,050 | 24,149 |
| 50–60 years | 78,903 | 54,331 | 41,848 | 46,433 | 43,886 | 48,270 |
| 60–70 years | 83,625 | 57,901 | 43,345 | 47,946 | 45,236 | 49,908 |
| 70–85 years | 133,235 | 98,062 | 66,593 | 69,881 | 65,801 | 68,759 |

**Supplementary Figure 1.**
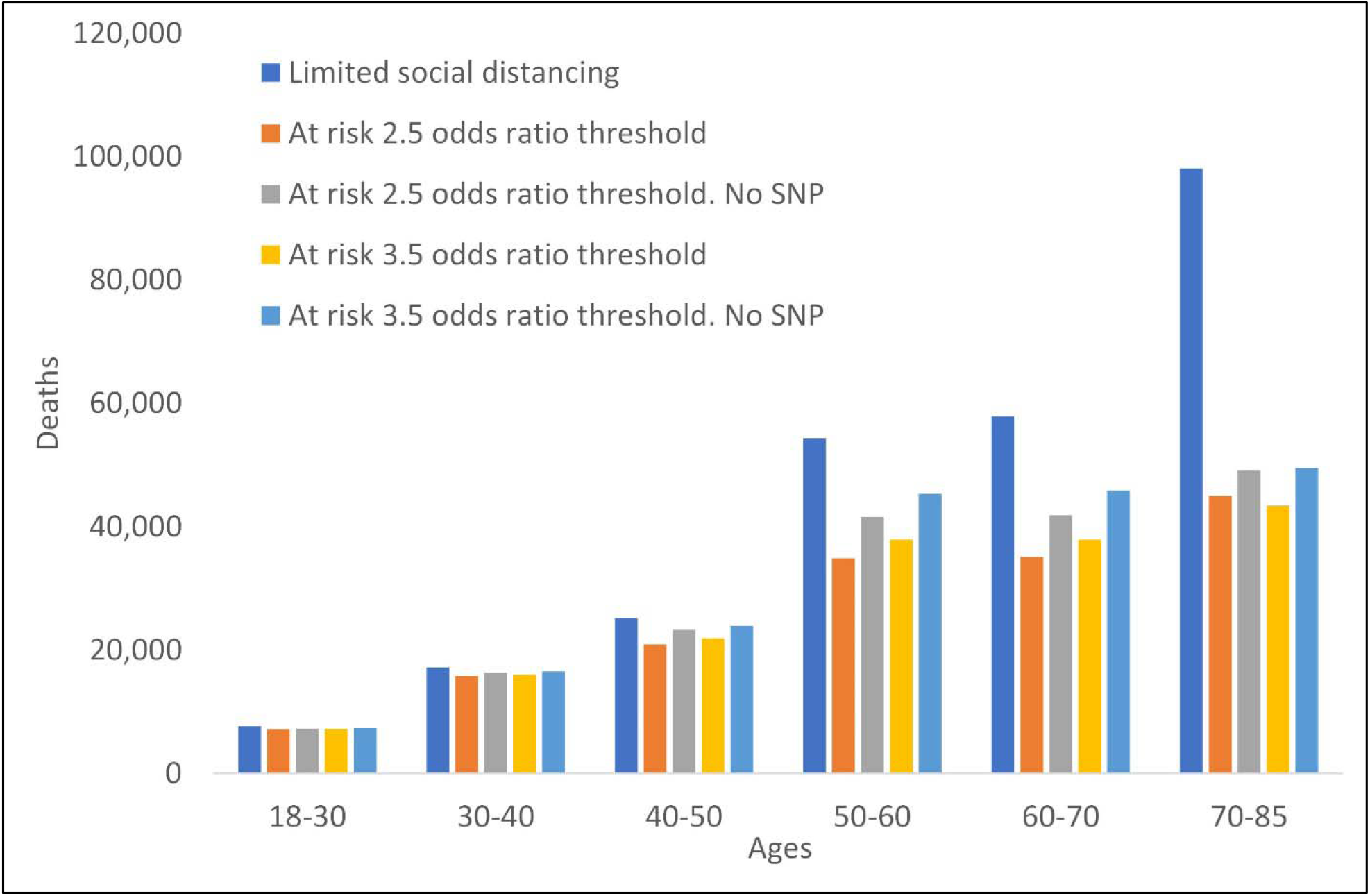

**Supplementary Figure 2.**
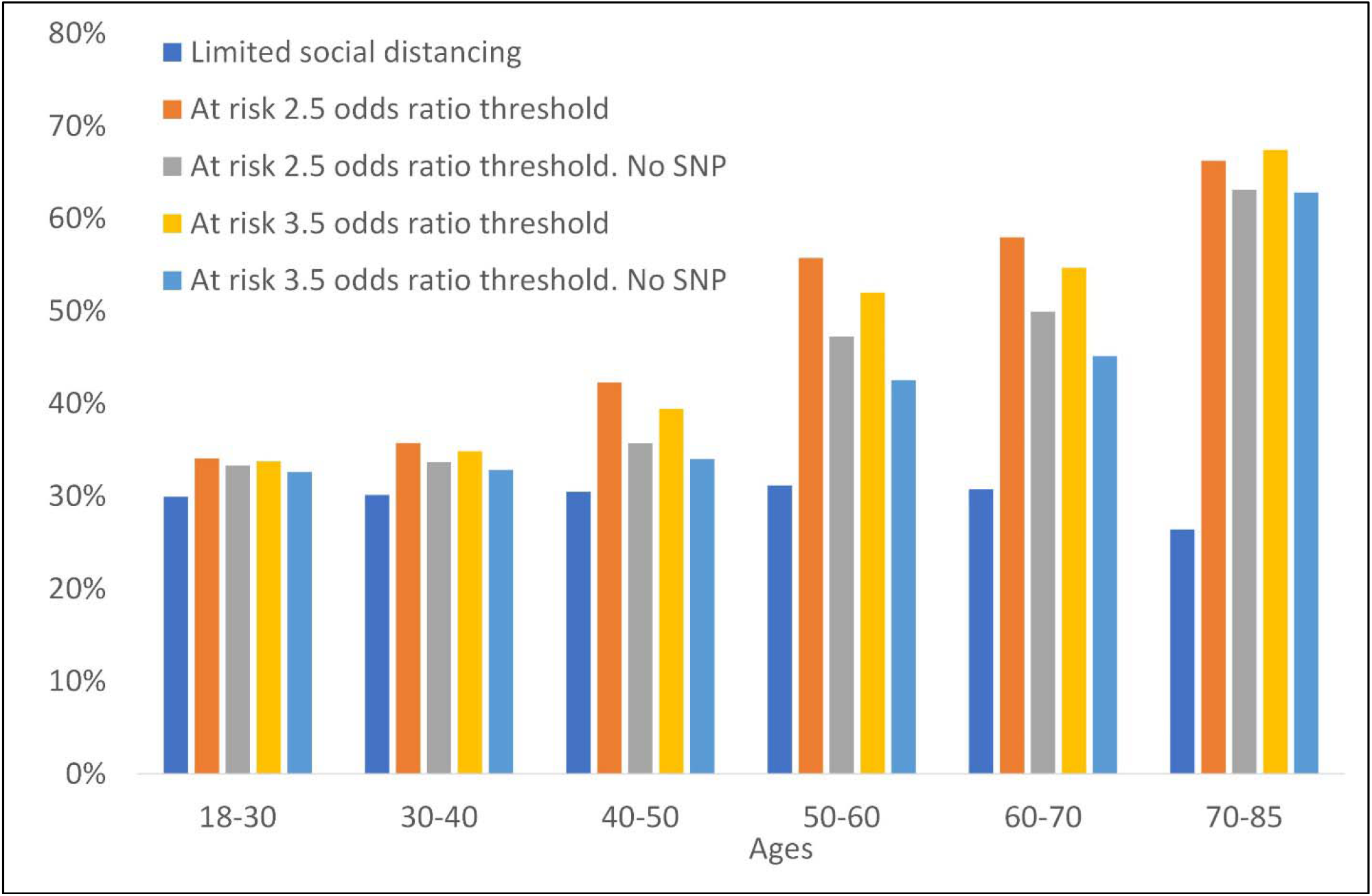

